# Education and Later-life Mortality: Evidence from a School Reform in Japan

**DOI:** 10.1101/2023.07.10.23292439

**Authors:** Kazuya Masuda, Hitoshi Shigeoka

## Abstract

We examine the mortality effects of a 1947 school reform in Japan, which extended compulsory schooling from primary to secondary school by as much as 3 years. The abolition of secondary school fees also indicates that those affected by the reform likely came from disadvantaged families who could have benefited the most from schooling. Even in this relatively favorable setting, we fail to find that the reform improved later-life mortality up to the age of 87 years, although it significantly increased years of schooling. This finding suggests limited health returns to schooling at the lower level of educational attainment.

## 1. Introduction

Economists argue that educated individuals live longer than uneducated people, because education expands access to resources, increases health investment, and improves health production efficiency through better knowledge and information gathering (Grossman 1972; Cutler and Lleras-Muney 2008). Consistent with this argument, a positive association between educational attainment and health status has been observed across time and space. However, whether this relationship is causal is still an ongoing question. Several studies have used reforms in compulsory schooling to estimate the causal impact of schooling on later-life mortality, but the evidence is highly mixed.^1^

We examine the impact of a 1947 school reform in Japan, which expanded compulsory schooling from primary to secondary schools, on mortality. We adopt two empirical approaches to estimate the impact of schooling on mortality. We first employ a regression discontinuity (RD) design at the monthly level to compare individuals born just before the school entry cutoff to those born just after, who were almost identical in age but were subject to school reform.

However, because students born immediately before and after the cutoff were also the oldest and youngest within their respective academic cohorts, we use the academic cohorts born before and after the school reform as the control group in a difference-in-regression discontinuity (Dif-in-RD) design to isolate the effect of additional schooling from that of relative age effects.

Using the 100% census in 2020, we demonstrate that the school reform led to significant increases in the completion rate of secondary school and, hence, years of schooling, by 0.12–0.21 years. However, we do not find that additional schooling reduces the mortality of the affected cohorts up to the age of 87 years. The mortality estimate is precisely 0, and we can rule out, with 95% confidence, that the school reform reduced mortality by more than 0.7 percentage points in the 50 years between 1971 and 2021, during which time as high as 67.2% of the affected cohorts died. Furthermore, we find no evidence of reductions in mortality from more specific causes of death or in the rates of outpatient visits and inpatient admissions, which could affect the quality of life. We also examine several socioeconomic outcomes available from the 1980 census. While the effect of the school reform on labor force participation and employment is very limited, the affected cohorts shift the industry from the primary sector to the tertiary sector, indicating more access to white-collar jobs. However, the magnitude of this shift is small.

Our results echo the null findings of education on mortality in the recent literature by Clark and Royer (2013) for the UK, Meghir et al. (2018) for Sweden, and Malamud et al. (2023) for Romania. Our study contributes to the widely studied, but unsettled, literature in several ways.

First, the characteristics of the affected cohorts (i.e., “compliers”) can be quite different from those in past studies. Since the pre-reform completion rate of secondary schools was already as high as 92% and the fee for secondary schools was abolished by the reform, those induced to study by the school reform were likely to come from disadvantaged families, most likely with financial constraints. Such children could benefit the most from improved labor market opportunities, knowledge acquired from additional schooling, and better peers.^2^ This situation sharply contrasts with other compulsory schooling reform settings, in which students— who would have dropped out of secondary education exactly at the minimum school-leaving age in the absence of the reform—are compelled to remain in school; hence, the monetary and health returns from more schooling could be small.

Second, the incremental years of schooling are as many as 3 years from grades 7 to 9, unlike most previous studies that mostly examine compulsory schooling extended by 1 year, such as the school reform in the UK. While the non-linear return on education by the initial level of education (e.g., primary schooling vs. college education) has been heavily discussed, potential non-linearity by a different incremental margin in years of schooling has received less attention in the literature.

Third, we can track the mortality of affected cohorts up to the age of 87 years, whereas other studies, at best, track mortality until ages in the early 70s. Clark and Royer (2013) for the UK, Meghir et al. (2018) for Sweden, and Malamud et al. (2023) for Romania track mortality up to the ages of 74, 75, and 71 years, respectively.^3^ An additional 10–15 years of follow-up period is crucial, as the mortality for the affected cohorts in our setting is 14% between the ages of 65 and 74 years, but as high as 31% between the ages of 75 and 84 years.

Finally, this is the first study on the causal estimate of schooling on later-life mortality outside the US and Europe, except for Taiwan, which expanded compulsory schooling in 1968 (Kan 2016), because the interventions need to have occurred many years ago to observe old-age mortality. Japan is ideal, as the school reform took place as early as 1947, the same year as that in the UK, which has been extensively studied (e.g., Oreopoulos 2006; Clark and Royer 2013).

Taken together, we examined the impact of education on mortality among individuals who might have benefited from additional schooling up to very late in their lives. Even in this favorable setting, we fail to find evidence of a positive return of schooling on mortality. While the discrepancy across studies could stem from different populations being affected by the reforms, our results show that the return of education on mortality is likely limited at the lower level of schooling.

## 2. Background

Prior to the postwar education reform, all pupils in Japan attended a 6-year common primary school from ages 6–12 years under the National School Order implemented in 1941. After completing sixth grade, post-primary education was not compulsory; they either opted out of the formal education system or continued schooling. There were three post-primary education tracks: general, academic, and vocational. Any pupils who wished to continue further studies, as long as they could cover school fees, attend a general-track 2-year secondary school. Alternatively, pupils who passed the exams could attend either an academic or vocational track school, both of which were recorded as “high school” (not “secondary school”) in the census in Japan.

Tuition for primary school was eliminated in 1900, but tuition for post-primary school was not. Each municipality determined the fee level for public schools. For example, a general-track secondary school charged 12 Yen per academic year in 1946 (Yubetsu-cho 1982), a large amount for a poor household when the average annual household income among the non-rich was approximately 237 Yen during the pre-war period (Minami et al. 1993).

In 1947, with the support of the General Headquarters, the Japanese government implemented a school reform, legally defined in the School Education Law (SEL). As all laws governing compulsory education were legislated at the national level, the timing of reform implementation was the same for all regions.

The main changes brought about by the reforms are as follows.

i. Compulsory full-time schooling was extended from 6 years (primary school) to 9 years (primary and secondary school).^4^
ii. The early selection of different tracks after grade six was abolished. Under the new system, all pupils were kept in a common general secondary school until the ninth grade.
iii. The fee for secondary school was abolished.

Note that two types of compliers were affected by the school reform. First, those who would have stopped at primary school in the absence of the law now completed secondary school under the new educational system. As the completion rate of secondary school, as shown in Section 5.1, was already as high as 84%–92% before the reform, room for improvement was limited. Nonetheless, their schooling increased by 3 years. Importantly, these compilers might have come from disadvantaged families who could not afford the fee before the reform. While we cannot directly examine the socioeconomics of compliers because of data limitations, these pupils could have benefited most from improving health knowledge or meeting more knowledgeable peers compared to the students who would have remained in school regardless of the law changes. By contrast, the fee had already been abolished in the UK at the time of its educational reform in 1947 (Oreopoulos 2006).

Second, those who would have stopped at secondary school without changes in the law now enjoyed an additional year of schooling, from 2 years of secondary school in the old system to 3 years in the new system. To the extent that students attended an additional year of secondary school owing to the abolition of school fees, they might have also come from relatively disadvantaged families.

The academic year in Japan starts in April, and Article 22 of the SEL obliges parents to send their children to primary school as soon as they turn 6 years of age before the next April. Consequently, children born between April and March comprise an academic cohort.

As the school reform applies to the cohort about to start secondary school in 1947, the cohort born after April 1934 (i.e., 12 years before the reform) was the first academic cohort directly affected by the reform. However, older academic cohorts, in particular those born in 1932 and 1933, at the end of the first and second year of the “old” secondary school at the time of reform, could be partially affected because the abolition of fees applied not only to entering students but also to current students. Although they were technically exempt from compulsory schooling, they may have decided to continue secondary school in the new system, probably because the remaining years of secondary education were free. Alternatively, they (or their parents) might have been afraid of being disadvantaged by competing with peers with more years of schooling in the future labor market.

One might think that students did not learn meaningfully in the turbulent environment immediately after WWII, but this does not seem to be supported.^5^ Figure A1 plots the student– teacher ratio (Panel A) and student–classroom ratio (Panel B) in secondary schools from 1943 to 1951. While the reform significantly increased secondary school enrollment, both measures remained approximately the same or improved slightly over time. Table A1 compares the Grade 7 curriculum (first year of secondary school) before and after the reform. Not surprisingly, nationalistic and militaristic curriculum content was removed and more meaningful subjects, such as reading, science, and foreign languages, were added. Finally, as shown in Section 5.4, we document some positive returns, albeit of a small magnitude, of education in the labor market. Indeed, nearly 90% of the cohorts just before the reform were already enrolled in secondary school, when it was not compulsory and required a fee, indicating that students (or their parents) saw some potential return from secondary education even at that time.

## 3. Data

Our primary sample consists of individuals born in Japan between April 1929 and March 1936 (96 birth year-month cohorts), including the three academic years before and after the affected cohorts (1932–1934).

### 3.1. Education

To estimate the impact of school reform on the level of completed education, we used the 100% 2020 Japanese census, which has become available recently. Notably, this census is the first in the history of Japanese census to distinguish between primary and secondary schools in the categories for the highest level of completed education.^6^ As a result, to the best of our knowledge, we are the first to document the impact of the 1947 school reform not only on educational attainment (“first stage”) but also on any reduced-form outcomes, such as labor and health outcomes.^7^ With more than 50,000 observations in each birth year-month cohort totaling 8 million observations in the sample period, we have substantial power to implement RD. The obvious downside is that the affected cohorts born between 1932 and 1934 were already 86–88 years at the time of the census. To the extent that individuals induced to attain more schooling come from disadvantaged families, they may die early and, hence, are not included in the census. This may attenuate the estimate of educational attainment.

Specifically, the 2020 census asked for the highest level of completed education in the following categories: primary school, secondary school, high school, junior college, college, and graduate education. We created a dummy variable for each level of educational completion. We also assigned a number of years of schooling to each level of education. As the census does not distinguish between old and new education systems, we used the pooled Social Stratification and Social Mobility Survey of 1985 and 1995, which separately recorded old 2-year and new 3-year secondary schools, along with birth year (not birth year-month) information, to impute years of schooling for those who had completed secondary school.^8^

Table A2 provides the imputed years of schooling across academic cohorts. It confirms that some fraction of the 1932 and 1933 academic cohorts, who are not subject to compulsory secondary education, completed secondary school in the new system, implying that we should expect discontinuities in the probability of secondary school completion at the April 1932 and 1933 cutoffs.

### 3.2. Mortality

We used vital statistics for 1971–2021 to estimate the impact of school reform on mortality. This administrative dataset, collected by the Ministry of Health, Labour and Welfare, captures all deaths in Japan, including the exact death dates and birth year-month of the deceased. January 1971 is the earliest year-month for which individual-level death records are available, and December 2021 is the latest available at the time of writing. Vital statistics contain the deaths of Japanese nationals that occurred outside Japan. Hence, migration bias is not a serious concern in our setting.

As the 2000 census is the first to report the birth year-month (not birth year-quarter), our running variable in RD, we used the 2000 census, which captures the population as of October 2000, as the starting point. We subtracted the cumulative deaths between January 1971 and October 2000 to construct the at-risk population for January 1971.^9^ Our main outcome is the mortality rate between 1971 and 2021, where the total deaths between January 1971 and December 2021 are divided by the population in January 1971 for each birth year-month cohort.^10^

In this way, we tracked each cohort for 50 years (ages from 37 to 87 years) for the 1932– 1934 birth cohorts. Approximately 59%–67% died between 1971 and 2021, conditional on survival up to 1971, providing a nearly complete picture of the impact of education on later-life mortality. For reference, the life expectancy in 2021 was 81.5 and 87.6 years for men and women, respectively.

One limitation is that individual-level mortality data were available only after 1971. Here, we tested for selective mortality up to 1971 using birth counts, the 2000 census, and deaths recorded between 1971 and 2000. Figure C1 shows no suggestive evidence of selective mortality at any relevant school entry cutoffs in the month of April.

Vital statistics also provide detailed information on the leading causes of death (ICD9 until 1994 and ICD10 after 1995), allowing us to examine the cause-specific mortality associated with health behaviors. We examined two leading causes of death, cancer and circulatory diseases, accounting for 33.7% and 24.3% of all deaths, respectively, in the 1932–1934 birth cohorts. In addition, we classified certain causes of death as preventable or treatable based on epidemiological literature, following Meghir et al. (2018) and Malamud et al. (2023). Preventable causes of death (e.g., accidents) should reflect risk-taking behaviors and health investments, whereas treatable causes of death (e.g., asthma) could be related to healthcare access.

## 4. Empirical strategy

We adopt two empirical methods: 1) RD with a linear spline to identify the effect on each academic cohort separately and 2) Dif-in-RD to identify the average effects of all affected cohorts. Throughout the study, we collapse the outcomes at the birth year-month level (denoted by *b*), as it is the level of our analysis in both designs.

### 4.1. Regression discontinuity design

Let *Y*_*b*_ be the outcome of interest for birth year-month cohort *b*. We run the following regression:

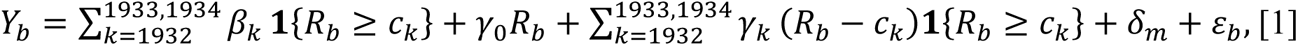

where *R*_*b*_ is the birth year-month cohorts, our running variable, which takes the value 1 in April 1929, 2 in May 1929, and 96 in March 1936.^11^ *c*_*k*_ (*k* = 1932, 1933, and 1934) are the April cutoffs for 1932, 1933, and 1934, respectively. **1**{*R*_*b*_ ≥ *c*_*k*_}, the first term in [1], takes one for cohorts born after each school entry cutoff, so each β_*k*_—our coefficient of interest— captures the *additional* effect of passing each April cutoff.

We allow each linear spline of running variable (*R*_*b*_ and (*R*_*b*_ − *c*_*k*_)**1**{*R*_*i*_ ≥ *c*_*k*_}, the second and third terms in [1]) to differ before the reform, for each of the three affected academic cohorts, and after the reform to capture the underlying relationship between the birth cohort and outcomes. δ_*m*_ is the calendar birth month FEs to address the relative age effects as the children born just after the school entry cutoff are the oldest within the respective grades (e.g., Bedard and Dhuey 2006; Black et al. 2011; Cascio and Schanzenbach 2016), and seasonality of births (Buckles and Hungerman 2013; Shigeoka 2015) can also have an independent effect on mortality. Here, we further interact each birth month FE with a dummy for cohorts born after the school reform to allow for different seasonality patterns across the pre– and post-reform (Clark and Royer 2013).

We estimate equation [1] by ordinary least squares using the number of observations in each cell (i.e., cohort size) as the weights. Heteroskedastic-robust standard errors, unless specified otherwise, are reported.

### 4.2. Difference-in-regression discontinuity design

While we account for relative age effects and seasonality of births by including birth month FEs in equation [1], we can also explicitly address it by using the academic cohorts born before and after the reform, as a comparison group (“control cohorts,” hereafter). This can be done by estimating an analogous regression model to equation [1] for the discontinuities of being born just after the cutoff in the control cohorts and then subtracting these discontinuities of the control cohorts from those of the treatment cohorts (Malamud et al. 2023).

We first organize each birth year-month cohort in relation to the month of April to form “synthetic” cohorts (i.e., from October of the previous year to September). Let *Q*_*b*_ be the *normalized* birth year-month cohorts in the range of –6 to 5, which take 0 in April, 1 in May, and 5 in September, and –1 in March, –2 in February, and –6 in the previous October. Then, we estimate the following “difference-in-discontinuities” model:

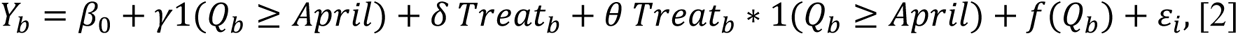

where 1(*Q*_*b*_ ≥ *Arril*) is an indicator of individuals born after April. *Treat*_*b*_ is an indicator equaling 1 for treated cohorts born during the 1932–1934 “synthetic” cohorts and 0 otherwise.^12^ *f*(*Q*_*b*_) now includes the interactions of our normalized running variable with both 1(*Q*_*b*_ ≥ *April*) and *Treat*_*b*_, allowing for different relationships between the outcome and birth year-month both before and after the school entry cutoff and in the treatment and control cohorts.

Thus, we flexibly control for an increasing trend in schooling and lower mortality for younger cohorts. Our coefficient of interest is the interaction term, θ, which estimates the impact of being born just after the cutoff among treatment cohorts over and above the effects in control cohorts before and after the school reform, which is captured by ɣ.^13^

The advantage of this approach is that it directly addresses relative age effects and provides the average effect for all three affected cohorts. The disadvantage is that running variable can take only 6 months on either side of the cutoffs by construction. To obtain balanced data around April, we remove 6 months from both ends of the sample used in equation [1], spanning October 1929 to September 1935 (or 84 birth year-month cohorts). The underlying assumptions for this empirical strategy are that (i) the relative age effects and seasonality are stable across the treatment and control cohorts and (ii) school cohort-specific shocks, other than the school reform, are balanced across the treatment and control cohorts (Grembi et al. 2016). We use this approach as a supplement to the RD specification.

We do not report the two-stage least squares estimates because the mortality data do not contain education, and there may be changes in school quality that are not captured by our imputed measure of schooling years. Instead, we follow Clark and Royer (2013), Meghir et al. (2018), and Malamud et al. (2023) and report the reduced-form estimates of school reform on mortality rather than split-sample instrumental variable estimates.

### 4.3. Balance tests

The key to our identification is that birth cohorts born before and after the relevant school entry cutoffs are comparable, except for changes in schooling. Panels A–C of Figure A1 plot the predetermined characteristics at birth: cohort size (i.e., density test by McCrary 2008), percentage of stillbirths, and sex ratio at birth. Except for strong seasonality, none of the three outcomes suddenly change at each cutoff. Table A3 shows that, once we control for calendar birth month FEs, most estimates at the cutoffs are neither statistically significant nor economically large.

## 5. Results

### 5.1. Effects on educational attainment

We first present graphical evidence of the first stage in Figure 1. Panel A plots the proportion of individuals who completed secondary school or higher between April 1929 and March 1937. We added vertical lines for April to determine each academic cohort. Each dot represents the average outcome for each birth year-month.

**Figure 1.**
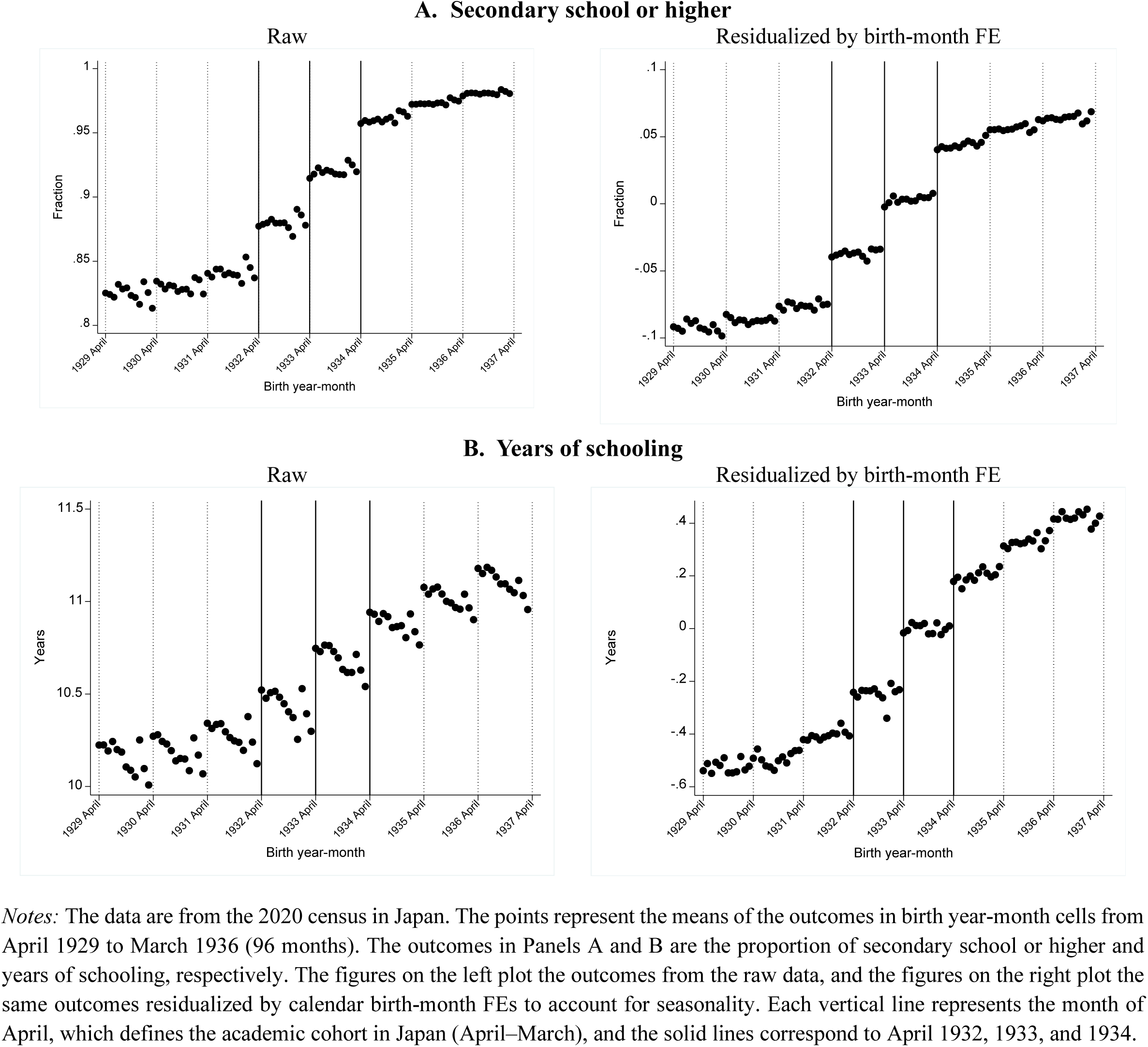
—Educational attainment.

The left graph of Panel A in Figure 1 shows that the proportion of individuals with secondary education or higher discontinuously increased in the Aprils of 1932, 1933, and 1934, with solid vertical lines. Not only the cohorts born after April 1934, who were directly affected by the reform, but also the cohorts born after April 1932 and 1933, who were at the end of the first and second year of secondary school at the time of reform, were also partially affected. In addition, the completion rate of secondary schools did not reach 100% immediately after the reform, posing the challenge of achieving universal coverage in a short period. To account for seasonality, the right graph of Panel A plots the outcome “residualized” by calendar birth month FEs. We are reassured that the results are similar. In addition, we do not observe similar discontinuities in other Aprils, which are used as control cohorts in Dif-in-RD.

Panel B of Figure 1 plots years of schooling as the outcome. The left graph shows that while seasonality is observed each April, the discontinuities at the relevant cutoffs are relatively larger than those of the surrounding cutoffs. The right graph, which partials out the birth month FEs, clearly shows large discontinuous jumps at the three relevant cutoffs, whereas the discontinuous jumps at the other cutoffs are mostly muted.

Table 1 reports a separate estimate at each of the three cutoffs from equation [1] in Panel A and a pooled estimate from equation [2] in Panel B. Column (1) of Panel A shows that the fraction of those who completed secondary school or higher increased by 2.8, 2.5 and 3.4 percentage points (p-value<0.01) at each 1932, 1933, and 1934 cutoff, off the mean of 0.84, 0.88, and 0.92, respectively. We find little spillover effect on higher education than on secondary school in columns (2) and (3), except for a 0.8-percentage point increase in college or above for the 1934 cutoff, which is the cohort directly affected by the reform.

**Table 1.** —Effects#on educational attainment.

|  | Secondary<br>or higher<br>(1) | High school<br>or higher<br>(2) | College<br>or higher<br>(3) | Years of<br>schooling<br>(4) |
| --- | --- | --- | --- | --- |
| **A. RD with multiple cutoffs** |  |  |  |  |
| 1932 cutoff | 0.028***<br>(0.002) | -0.011***<br>(0.002) | 0.001<br>(0.001) | 0.117***<br>(0.012) |
| 1933 cutoff | 0.025***<br>(0.002) | -0.004<br>(0.003) | 0.001<br>(0.002) | 0.209***<br>(0.019) |
| 1934 cutoff | 0.034***<br>(0.003) | -0.011*<br>(0.006) | 0.008***<br>(0.002) | 0.178***<br>(0.035) |
| Mean below 1932 cutoff | 0.842 | 0.550 | 0.073 | 10.270 |
| Mean below 1933 cutoff | 0.881 | 0.539 | 0.078 | 10.431 |
| Mean below 1934 cutoff | 0.921 | 0.527 | 0.080 | 10.673 |
| N of birth year-month | 96 | 96 | 96 | 96 |
| N of observations | 4,818,576 | 4,818,576 | 4,818,576 | 4,818,576 |
| **B. Dif-in-RD (pooled cutoffs)** |  |  |  |  |
| 1(After April) | 0.007***<br>(0.002) | 0.049***<br>(0.006) | 0.015***<br>(0.002) | 0.248***<br>(0.029) |
| 1(After April)×Treated | 0.026***<br>(0.004) | -0.020**<br>(0.010) | -0.001<br>(0.003) | 0.111**<br>(0.050) |
| Mean | 0.884 | 0.529 | 0.075 | 10.431 |
| N of birth year-month | 84 | 84 | 84 | 84 |
| N of observations | 4,204,000 | 4,204,000 | 4,204,000 | 4,204,000 |

*Notes:* The data are from the 2020 census in Japan, collapsed at birth year-month cells from April 1929 to March 1936 (96 months) in Panel A and from October 1929 to September 1935 (84 months) in Panel B. Panel A reports the estimates from equation [1], with robust standard errors in parentheses. The 1932, 1933, and 1934 cutoffs are dummies that take 1 when the cohort is above April each year. Panel B reports the estimates from equation [2] with robust standard errors in parentheses. 1(After April) is an indicator equaling 1 for individuals born after April and 0 otherwise. Treated is an indicator equaling 1 for treated cohorts born between October 1932 and September 1935 and 0 otherwise. See Table A1 to assign the years of schooling to each birth cohort in column (4). Significance levels:
\*\*\* p<0.01, \*\* p<0.05, \* p<0.10.

Column (4) of Table 1 reports that years of schooling increased by 0.12, 0.21, and 0.18 at the 1932, 1933, and 1934 cutoffs, respectively. As mentioned in Section 2, such increased schooling years reflect both 1) the higher completion rate of secondary school among those who would have stopped at primary school and 2) a 1-year extension of secondary schooling for those who would have stopped at secondary school without the law change. We compute the contribution of the former by multiplying the estimates of secondary school or higher in column (1) by 3 years (as little spillover is observed) and dividing it by the estimate of years of schooling in column (4). The former accounted for 71.8%, 35.9%, and 57.3% of the increase in schooling years at the 1932, 1933, and 1934 cutoffs, respectively.

Panel B of Table 1 reports the Dif-in-RD estimates, where we also report the ɣɣ, which captures the relative age effects for the control cohorts, along with θθ, our coefficient of interest for the interaction term in equation [2]. Column (1) shows that the fraction of completing secondary school or higher increases by 2.6 percentage points, and this translates into 0.11 years of additional schooling on average in column (4). Figure B1 visually shows these relationships by plotting the outcomes for the treatment cohorts (first row), control cohorts (second row), and their differences (third row) around April, normalized to 0. While we observe a sharp increase in both secondary school or above and years of schooling at the cutoff for the treated cohorts, we do not see similar jumps for the control cohorts, resulting in positive Dif-in-RD estimates.

We compare the magnitude of educational attainment with the past literature that also exploits changes in compulsory schooling in the RD framework. Our estimates of 0.12–0.21 additional years of schooling are much smaller than those of Clark and Royer (2013), with 0.45 and 0.35 years in 1947 and 1972 reforms in the UK, smaller or close to those of Meghir et al. (2018) with 0.25 years in Sweden, but larger than those of Malamud et al. (2023) with 0.12 years in Romania. In contrast to prior studies where school reforms affected a large fraction of cohorts, such as in the UK, making them average population effects, the reform in this study affected a relatively small fraction of students who could benefit from additional schooling.

Figure B2 and Table B1 show heterogeneity by gender. We find similar impact sizes on the completion rate of secondary school or higher for both genders.^14^

### 5.2. Effects on mortality

We now examine the impact of school reform on mortality, which is our main outcome. Figure 2 plots all-cause mortality in Panel A, followed by cancer and circulatory diseases in Panels B and C, and preventable or treatable causes of death in Panels D and E. The negative slopes in all graphs reflect that people are younger along the horizontal axis. However, we find no large or significant breaks in this trend at the relevant cutoffs, providing visual evidence that the reform did not affect mortality. Figure C2 plots all-cause mortality for treatment cohorts, control cohorts, and their differences around April in the spirit of Dif-in-RD; no obvious discontinuities are observed at the cutoffs for any of the three plots.

**Figure 2.**
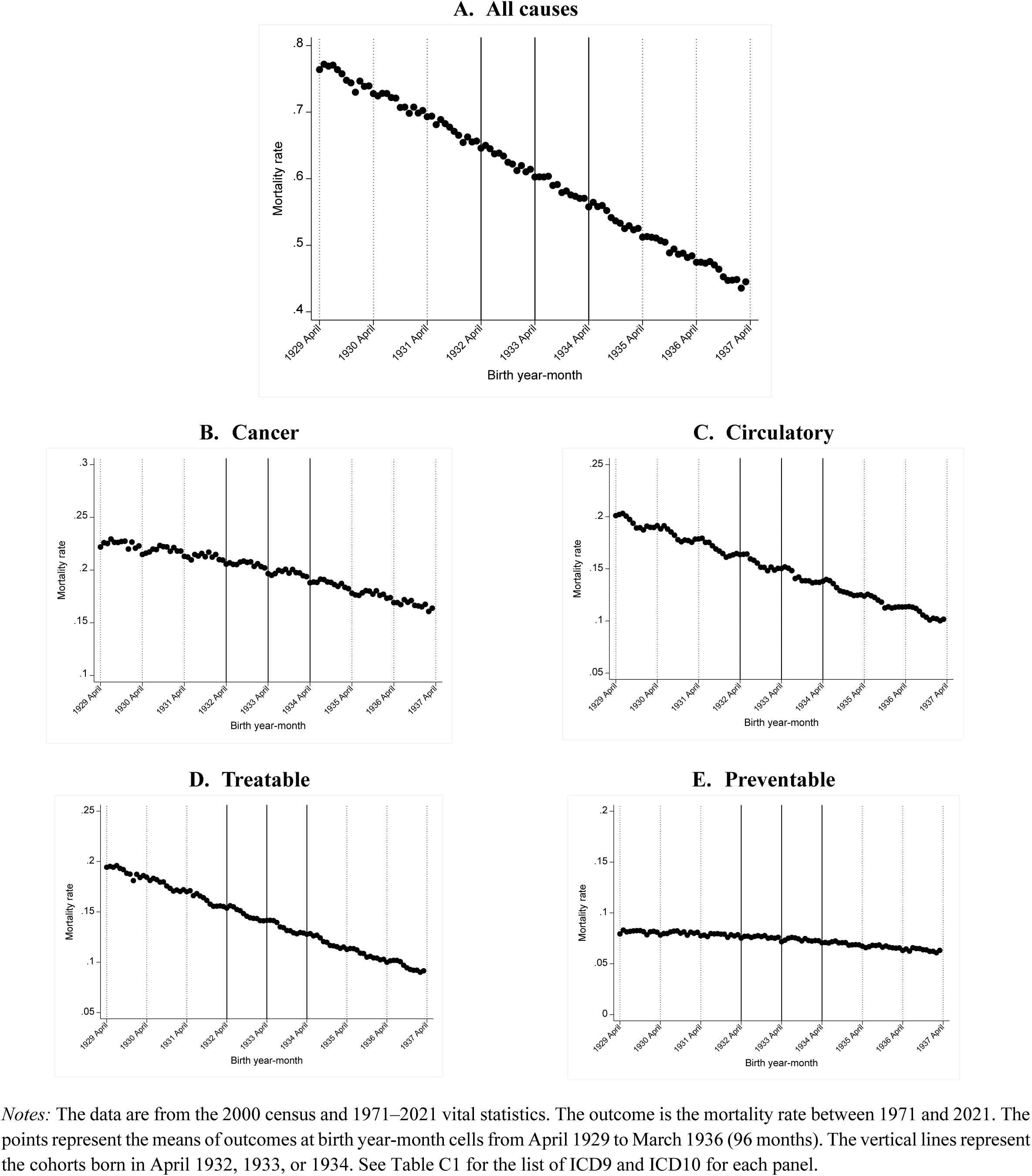
—Mortality.

Table 2 reports the estimates from RD in Panel A and Dif-in-RD in Panel B. Column (1) for all-cause mortality shows that the mortality estimates are far from statistically significant. The estimates for the 1932, 1933, and 1934 cutoffs are –0.0025, 0.0002, and 0.0005, respectively, from the mean of 0.672, 0.628, and 0.585, respectively. While the natural question is about how much statistical power we have to detect the mortality effect, our reduced-form estimate of mortality is precisely close to 0. For example, the lower bound of the 95% CI of the 1932 cutoff estimate, the largest among all three estimates, is –0.74 percentage points, which is just a 1.1% reduction from the mean mortality rate of 67.2% between 1971 and 2021. The rest of the table for cancer and circulatory diseases (columns 2 and 3) and preventable or treatable causes of death (columns 4 and 5) show little change in mortality at each threshold.

**Table 2.** —Effects on mortality.

|  | All causes | Cause of deaths |  |  |  |
| --- | --- | --- | --- | --- | --- |
|  |  | Cancer | Circulatory diseases | Treatable | Preventable |
|  | (1) | (2) | (3) | (4) | (5) |
| **A. RD with multiple cutoffs** |  |  |  |  |  |
| 1932 cutoff | -0.0025<br>(0.0025) | 0.0002<br>(0.0011) | -0.0012<br>(0.0009) | 0.0001<br>(0.0011) | -0.0001<br>(0.0006) |
| 1933 cutoff | 0.0002<br>(0.0027) | -0.0000<br>(0.0013) | -0.0007<br>(0.0013) | 0.0011<br>(0.0014) | 0.0001<br>(0.0008) |
| 1934 cutoff | 0.0005<br>(0.0038) | -0.0009<br>(0.0014) | 0.0015<br>(0.0015) | 0.0013<br>(0.0012) | 0.0007<br>(0.0009) |
| Mean below 1932 cutoff | 0.672 | 0.213 | 0.169 | 0.161 | 0.078 |
| Mean below 1933 cutoff | 0.628 | 0.206 | 0.155 | 0.147 | 0.076 |
| Mean below 1934 cutoff | 0.585 | 0.197 | 0.142 | 0.134 | 0.074 |
| N of birth year-month | 96 | 96 | 96 | 96 | 96 |
| N of observations | 13,237,894 | 13,237,894 | 13,237,894 | 13,237,894 | 13,237,894 |
| **B. Dif-in-RD (pooled cutoffs)** |  |  |  |  |  |
| 1(After April) | -0.0071***<br>(0.0016) | -0.0054***<br>(0.0008) | 0.0019***<br>(0.0008) | 0.0001<br>(0.0008) | -0.0022***<br>(0.0004) |
| 1(After April)×Treated | 0.0033<br>(0.0026) | 0.0006<br>(0.0011) | 0.0013<br>(0.0013) | 0.0020*<br>(0.0011) | 0.0002<br>(0.0007) |
| Mean | 0.617 | 0.205 | 0.152 | 0.143 | 0.076 |
| N of birth year-month | 84 | 84 | 84 | 84 | 84 |
| N of observations | 11,624,394 | 11,624,394 | 11,624,394 | 11,624,394 | 11,624,394 |

*Notes:* The data are from the 2000 census, and 1971–2021 vital statistics collapsed at birth year-month cells from April 1929 to March 1936 (96 months) in Panel A, and October 1929 to September 1935 (84 months) in Panel B. Panel A reports the estimates from equation [1], with robust standard errors in parentheses. The 1932, 1933, and 1934 cutoffs are dummies that take 1 when the cohort is above April each year. Panel B reports the estimates from equation [2] with robust standard errors in parentheses. 1(After April) is an indicator equaling 1 for individuals born after April and 0 otherwise. Treated is an indicator equaling 1 for treated cohorts born between October 1932 and September 1935 and 0 otherwise. See Table C1 for the list of ICD9 and ICD10 for each cause of death. The number of observations reflects the number of survivors up to January 1971, our population at risk. Significance levels: \*\*\* p<0.01, \*\* p<0.05, \* p<0.10.

*Heterogeneity*—. Figure C3 and Table C2 examine heterogeneity by gender, as men and women are expected to follow different underlying health processes as they age. We did not find any effects on overall or cause-specific mortality in either gender. Table C3 divides the sample by period (1972–1999 vs. 2000–2021) and age (50–59, 60–69, and 70–79 years); however, we do not observe discernable patterns in all-cause mortality.

### 5.3. Effects on healthcare utilization

Appendix Section D examines the effect of school reform on outpatient visits and inpatient admissions, which may affect the quality of life, but not necessarily its length. See Appendix Section E for the dataset used in this analysis, the Patient Survey, also used by Shigeoka (2014). Figures D1 and D2 show no meaningful breaks in the utilization outcomes at any relevant cutoff. Table D1 confirms the visual patterns.

### 5.4. Effects on labor market outcomes

The primary focus of this study is to examine the effects of school reform on mortality. Nonetheless, examining whether the expansion affected other relevant outcomes helps us understand the underlying mechanisms between education and mortality.

We examine several labor market outcomes in the 1980 census, which was the earliest census available at the individual level. The 1932–1934 birth cohorts were aged 46–48 years in 1980; thus, they were still in the labor force, as the mandatory retirement age was 60 years. Note that birth is reported only at the quarter level in 1980, and thus, we estimate the variant of equation [1] where the birth year-month, our running variable, is replaced by birth year-quarter.

Table 3 reports the results. Columns (1) and (2) detect no changes in labor force participation or employment at any cutoffs, both of which are already quite high and thus have little room for improvement. Columns (3)–(5), which examine the choice of industry sectors, reveal that the affected cohorts shift the industry from primary sector to tertiary sector, indicating increased access to white-collar jobs. However, the magnitude is relatively small compared to the mean. For example, the fraction of those who work in the tertiary industry increased by 1.2 percentage points at the 1934 cutoff, off the mean of 0.50 or 2.4%. Finally, column (6) indicates that the proportion of individuals with high-skill jobs is slightly higher at some cutoffs, but the magnitudes are relatively small again.^15^ Unfortunately, the Japanese census lacks income or earnings.

**Table 3.** —Effects on labor market outcomes.

|  | Labor force<br>participation | Employment | Work in |  |  | High skill<br>job |
| --- | --- | --- | --- | --- | --- | --- |
|  |  |  | primary<br>sector | secondary<br>sector | tertiary<br>sector |  |
|  | (1) | (2) | (3) | (4) | (5) | (6) |
| **RD with multiple cutoffs** |  |  |  |  |  |  |
| 1932 cutoff | 0.001<br>(0.002) | 0.001<br>(0.002) | -0.001<br>(0.001) | -0.004*<br>(0.002) | 0.005**<br>(0.002) | -0.002**<br>(0.001) |
| 1933 cutoff | 0.003<br>(0.002) | 0.003<br>(0.002) | -0.004***<br>(0.001) | -0.001<br>(0.004) | 0.005<br>(0.003) | 0.000<br>(0.002) |
| 1934 cutoff | 0.002<br>(0.002) | 0.002<br>(0.002) | -0.008***<br>(0.003) | -0.004<br>(0.003) | 0.012***<br>(0.002) | 0.005**<br>(0.002) |
| Mean below 1932 cutoff | 0.797 | 0.785 | 0.142 | 0.364 | 0.493 | 0.146 |
| Mean below 1933 cutoff | 0.801 | 0.790 | 0.133 | 0.370 | 0.497 | 0.140 |
| Mean below 1934 cutoff | 0.803 | 0.791 | 0.123 | 0.376 | 0.500 | 0.137 |
| N of birth year-quarter | 32 | 32 | 32 | 32 | 32 | 32 |
| N of observations | 12,823,297 | 12,823,297 | 10,121,690 | 10,121,690 | 10,121,690 | 10,121,690 |

*Notes:* The data are from the 1980 census in Japan collapsed birth year-quarter cells from April 1929 to March 1936 (96 months). The table reports the estimates from equation [1], where birth year-month is replaced by birth year-quarter, with robust standard errors in parentheses. The 1932, 1933, and 1934 cutoffs are dummies that take 1 when the cohort is above April each year. The high-skill dummy is defined as those who work as professionals or managers, following ILO standards. Significance levels: \*\*\* p<0.01, \*\* p<0.05, \* p<0.10.

In summary, while we find that the affected cohorts seem to obtain better jobs, the magnitude of improvement in labor market outcomes seems limited. It is possible that, as nearly 90% of academic peers had already completed secondary schooling before the reform, the signaling effect of secondary education was limited.

Indeed, the effects of compulsory schooling on labor market outcomes in similar settings are highly mixed. While Oreopoulos (2006) finds that the 1947 UK reform raised income, Devereux and Hart (2010) and Clark (2023) find much smaller or zero effects. Similarly, Meghir and Palme (2005) find that the 1948 Swedish reform led to only a slight increase in income for men (2%), but no effect for women. Furthermore, Malamud et al. (2023) find a limited return of education on most labor market outcomes in Romania.

## 6. Conclusion

We examine the extent to which the positive and well-documented correlation between education and mortality is causal. We exploit sharp across-cohort differences in educational attainment due to Japan’s 1947 school reform, which extended compulsory schooling from 6 to 9 years. The abolition of school fees by the reform indicates that compliers likely came from disadvantaged families who could have benefited the most from schooling. Even in this relatively desirable setting, we fail to find that the affected cohorts improve later-life mortality up to the age of 87 years. This finding suggests that health returns to schooling are limited, at least at the lower level of educational attainment.

In terms of external validity, while Japan’s high-quality universal insurance system, implemented in 1961 (Kondo and Shigeoka 2013), has potential to reduce the education-health gradient, financial resources could still improve health outcomes, as people have to pay for coinsurance. Furthermore, the other benefits of education, such as better knowledge and better peers, should still be present.

We should emphasize that this result does not imply that the education return on mortality is insignificant throughout the education distribution. Some studies document that health returns from college education can be positive (Buckles et al. 2016; Connolly 2021; Fletcher and Noghanibehambari 2021). More research is warranted to understand whether such discrepancies in the findings arise from the various roles of schooling in improving health, differential complier characteristics, or a combination of both, at a different level of education attainment.

## Data Availability

Mortality, outpatient and inpatient data can be accessed through the Ministry of Health, Labour and Welfare in Japan.

## Online Appendix (Not for Publication)

## Appendix A: Other figures and tables

**Figure A1.**
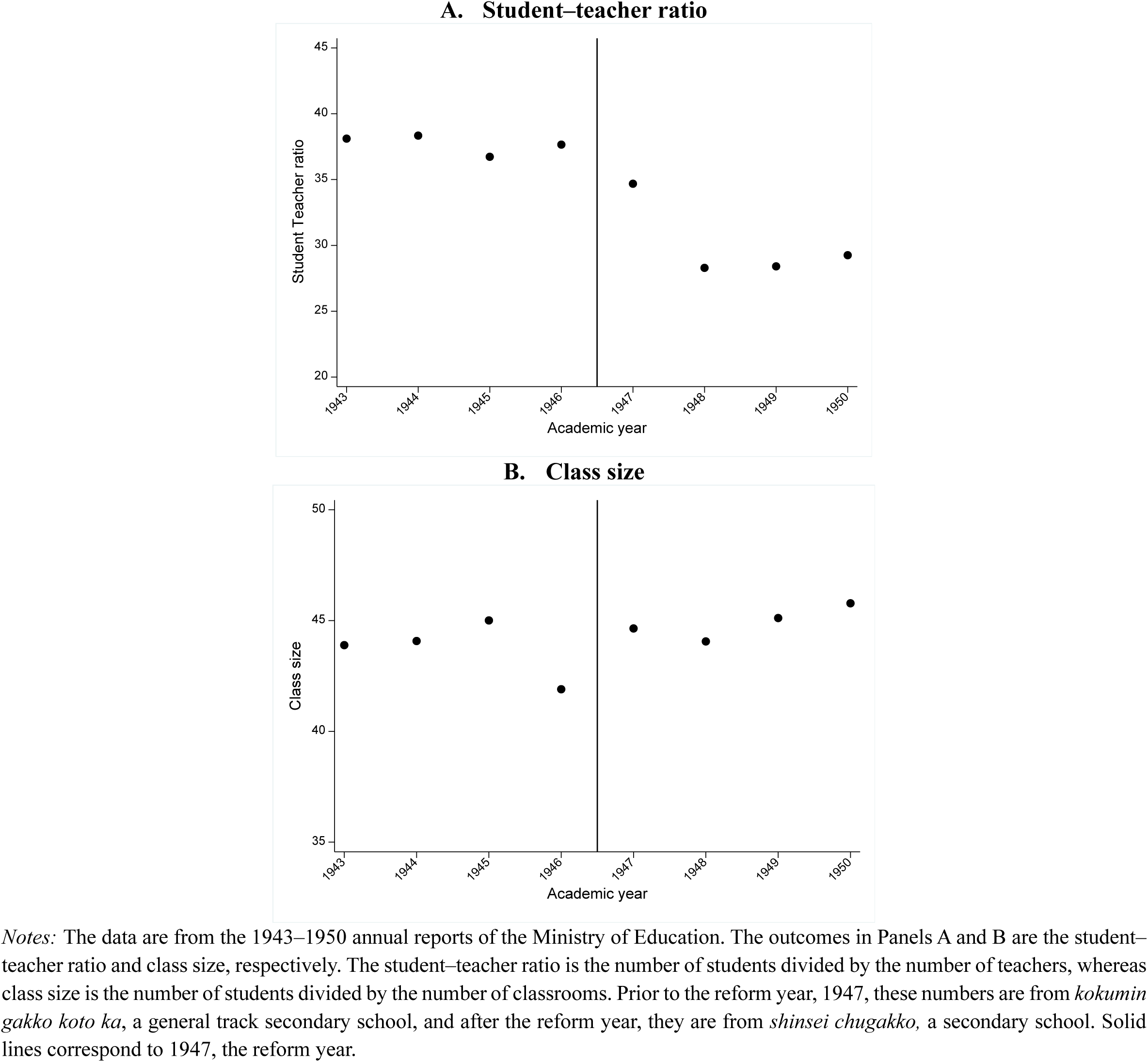
—Quality of secondary education.

**Figure A2.**
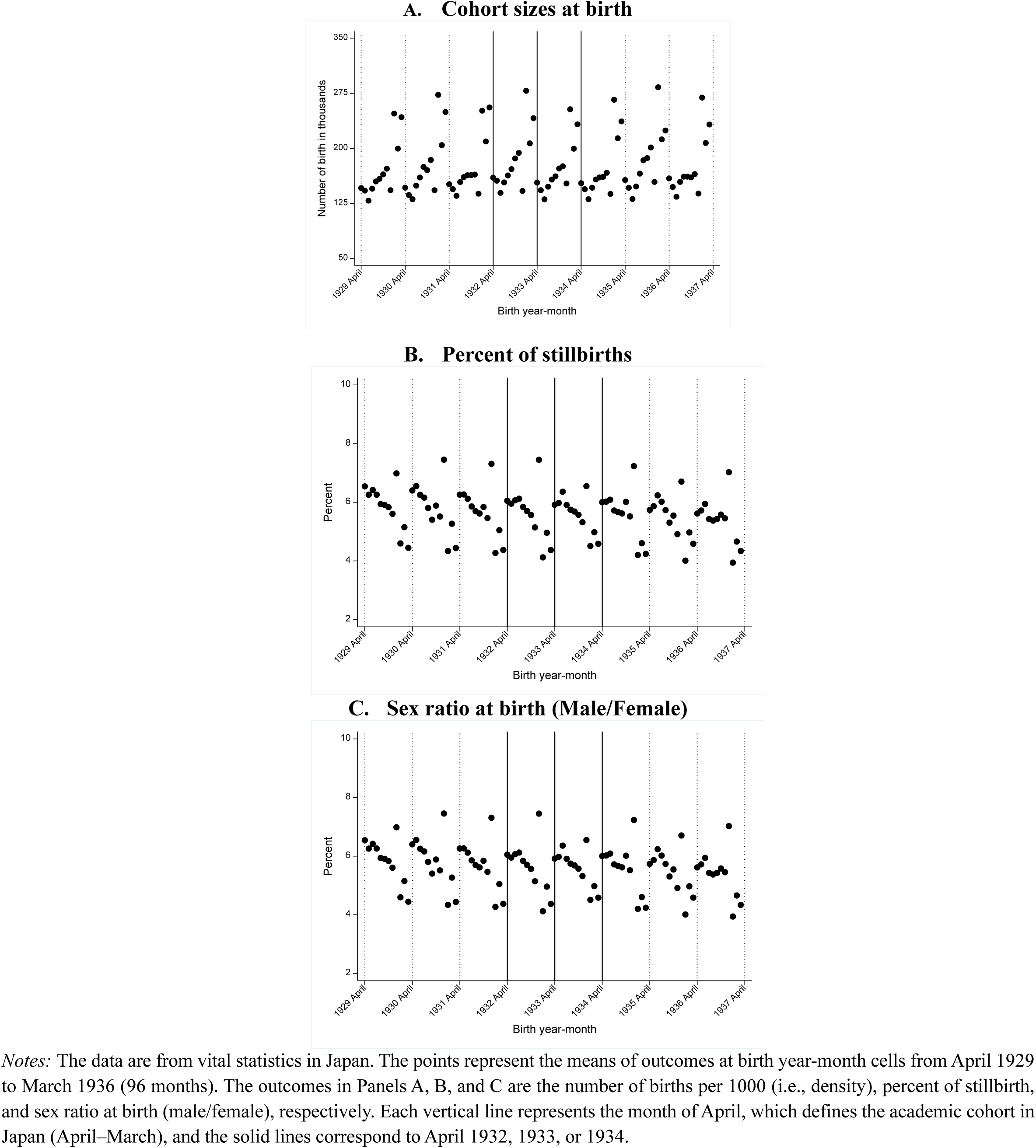
—Predetermined outcomes.

**Table A1.**
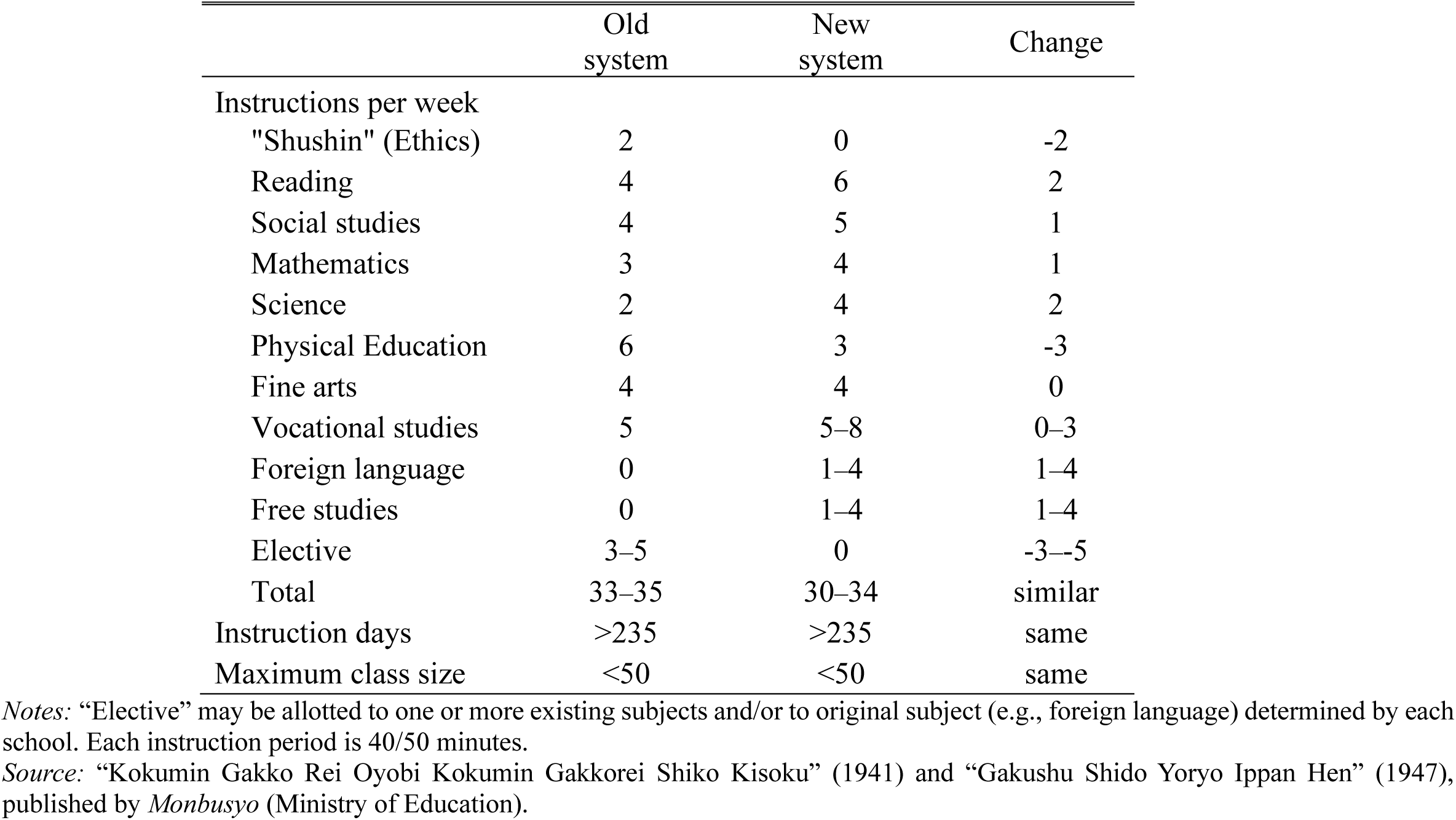
—Curriculum of grade 7 before and after the school reform.

**Table A2.** —Assigned years of schooling for secondary school completion.

| birth year | Old system<br>(8 years) | New system<br>(9 years) | Assigned years of schooling |
| --- | --- | --- | --- |
| 1930 | 100.0% | 0.0% | 8.00 |
| 1931 | 91.9% | 8.1% | 8.08 |
| 1932 | 62.3% | 37.7% | 8.38 |
| 1933 | 17.1% | 82.9% | 8.83 |
| 1934 | 0.0% | 100.0% | 9.00 |

*Notes:* The data are from 1985 and 1995 from Social Stratification and Social Mobility (SSM). See Appendix E for details of the dataset.

**Table A3.** —Balance test.

|  | Cohort size at<br>births<br>(in thousands) | Percent of<br>stillbirth | Sex ratio<br>at birth |
| --- | --- | --- | --- |
|  | (1) | (2) | (3) |
| **A. RD with multiple cutoffs** |  |  |  |
| 1932 cutoff | 7.087*<br>(3.753) | 0.028<br>(0.094) | 0.006<br>(0.007) |
| 1933 cutoff | -4.720<br>(5.108) | 0.010<br>(0.128) | 0.018*<br>(0.010) |
| 1934 cutoff | 5.792<br>(4.999) | -0.073<br>(0.153) | 0.005<br>(0.018) |
| Mean below 1932 cutoff | 174.373 | 5.677 | 1.049 |
| Mean below 1933 cutoff | 182.718 | 5.612 | 1.050 |
| Mean below 1934 cutoff | 173.365 | 5.592 | 1.057 |
| N of birth year-month | 96 | 96 | 96 |
| N of observations | - | 16,893,206 | 16,893,206 |
| **B. Dif-in-RD (pooled cutoffs)** |  |  |  |
| 1(After April) | -114.627***<br>(11.809) | 2.049***<br>(0.199) | 0.061***<br>(0.008) |
| 1(After April)×Treated | 4.097<br>(17.402) | -0.165<br>(0.268) | 0.005<br>(0.014) |
| Mean | 200.950 | 5.302 | 1.046 |
| N of birth year-month | 84 | 84 | 84 |
| N of observations | - | 14,844,291 | 14,844,291 |

*Notes:* The data are from vital statistics in Japan collapsed at birth year-month cells from April 1929 to March 1936 (96 months) in Panel A and from October 1929 to September 1935 (84 months) in Panel B. Panel A reports the estimates from equation [1], with robust standard errors in parentheses. The 1932, 1933, and 1934 cutoffs are dummies that take a value of 1 when the cohort is above April each year. Panel B reports the estimates from equation [2] with robust standard errors in parentheses. 1(After April) is an indicator equaling 1 for individuals born after April and 0 otherwise. Treated is an indicator equaling 1 for treated cohorts born between October 1932 and September 1935 and 0 otherwise. Significance levels: \*\*\* p<0.01, \*\* p<0.05, \* p<0.10.

## Appendix B: Results on educational attainment

**Figure B1.**
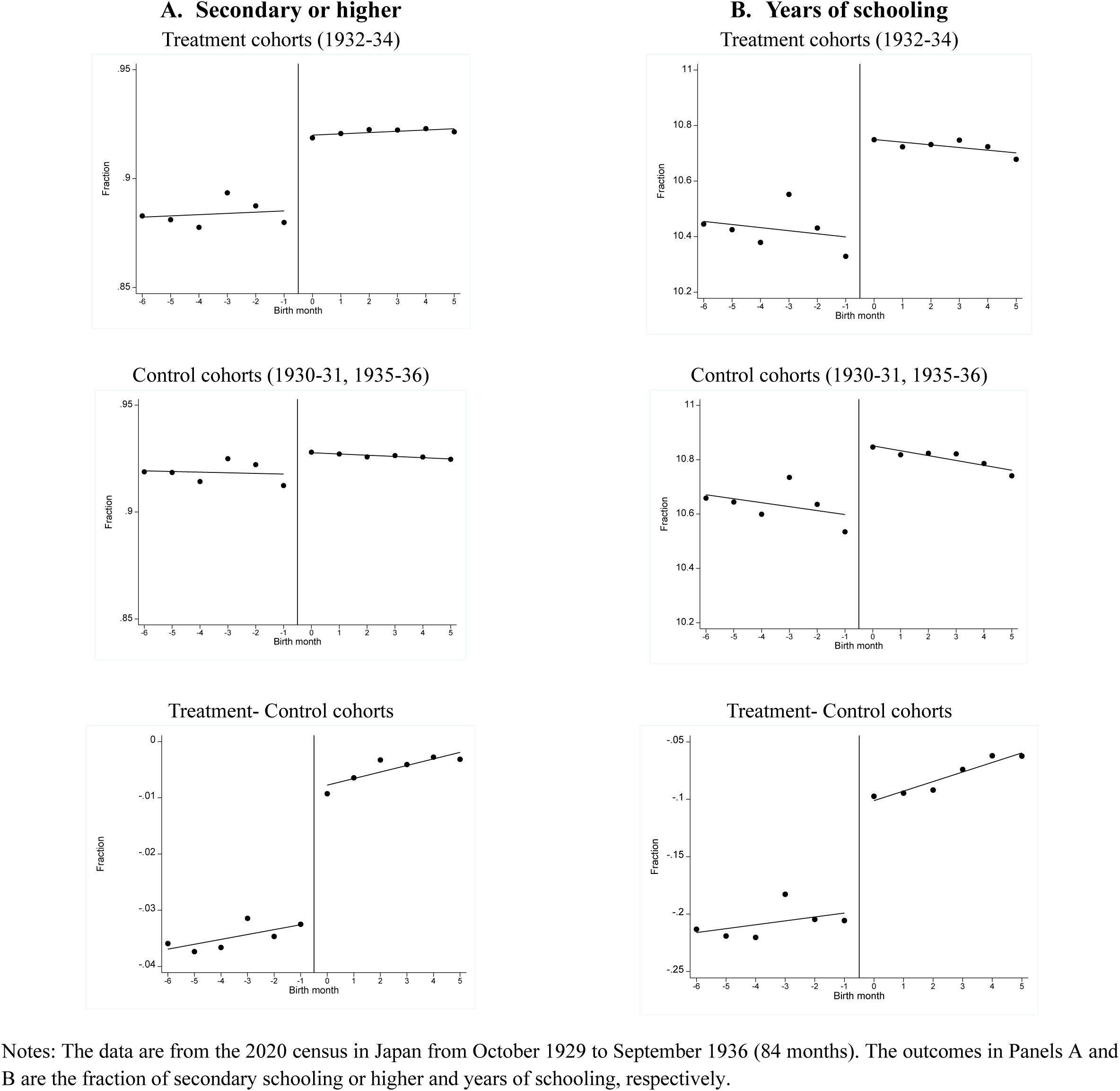
—Educational attainment by Dif-in-RD.

**Figure B2.**
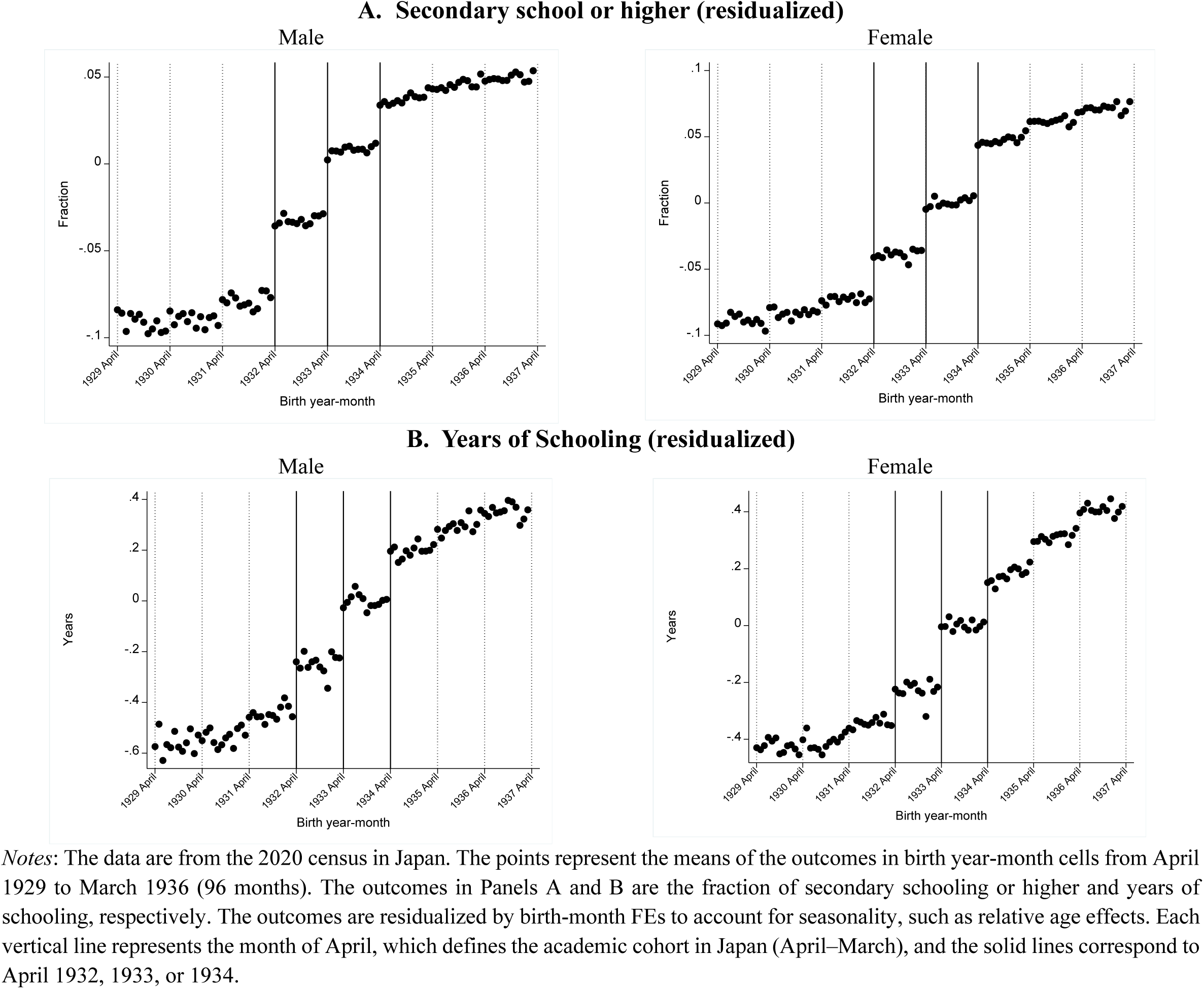
—Educational attainment by gender.

**Table B1.** —Educational attainment by gender.

|  | Secondary or higher |  | Years of schooling |  |
| --- | --- | --- | --- | --- |
|  | Male | Female | Male | Female |
|  | (1) | (2) | (3) | (4) |
| **A. RD with multiple cutoffs** |  |  |  |  |
| 1932 cutoff | 0.036***<br>(0.002) | 0.023***<br>(0.002) | 0.157***<br>(0.021) | 0.085***<br>(0.014) |
| 1933 cutoff | 0.026***<br>(0.002) | 0.023***<br>(0.003) | 0.213***<br>(0.030) | 0.189***<br>(0.022) |
| 1934 cutoff | 0.025***<br>(0.003) | 0.039***<br>(0.003) | 0.218***<br>(0.040) | 0.138***<br>(0.028) |
| Mean below 1932 cutoff | 0.861 | 0.846 | 10.825 | 9.994 |
| Mean below 1933 cutoff | 0.907 | 0.886 | 11.022 | 10.109 |
| Mean below 1934 cutoff | 0.948 | 0.926 | 11.268 | 10.339 |
| N of birth year-month | 96 | 96 | 96 | 96 |
| N of observations | 1,774,317 | 3,044,259 | 1,774,317 | 3,044,259 |
| **B. Dif-in-RD (pooled cutoffs)** |  |  |  |  |
| 1(After April) | 0.005***<br>(0.002) | 0.008***<br>(0.003) | 0.314***<br>(0.031) | 0.192***<br>(0.029) |
| 1(After April)×Treated | 0.027***<br>(0.004) | 0.025***<br>(0.005) | 0.149***<br>(0.056) | 0.088*<br>(0.051) |
| Mean | 0.905 | 0.867 | 10.957 | 10.108 |
| N of birth year-month | 84 | 84 | 84 | 84 |
| N of observations | 1,539,314 | 2,664,686 | 1,539,314 | 2,664,686 |

*Notes:* The data are from the 2020 census in Japan, collapsed at birth year-month cells from April 1929 to March 1936 (96 months) in Panel A and from October 1929 to September 1935 (84 months) in Panel B. Panel A reports the estimates from equation [1], with robust standard errors in parentheses. The 1932, 1933, and 1934 cutoffs are dummies that take 1 when the cohort is above April each year. Panel B reports the estimates from equation [2] with robust standard errors in parentheses. 1(After April) is an indicator equaling 1 for individuals born after April and 0 otherwise. Treated is an indicator equaling 1 for treated cohorts born between October 1932 and September 1935 and 0 otherwise. See Table A1 to assign years of schooling to each birth cohort in columns (3) and (4). Significance levels: \*\*\* p<0.01, \*\* p<0.05, \* p<0.10.

## Appendix C: Results on mortality

**Figure C1.**
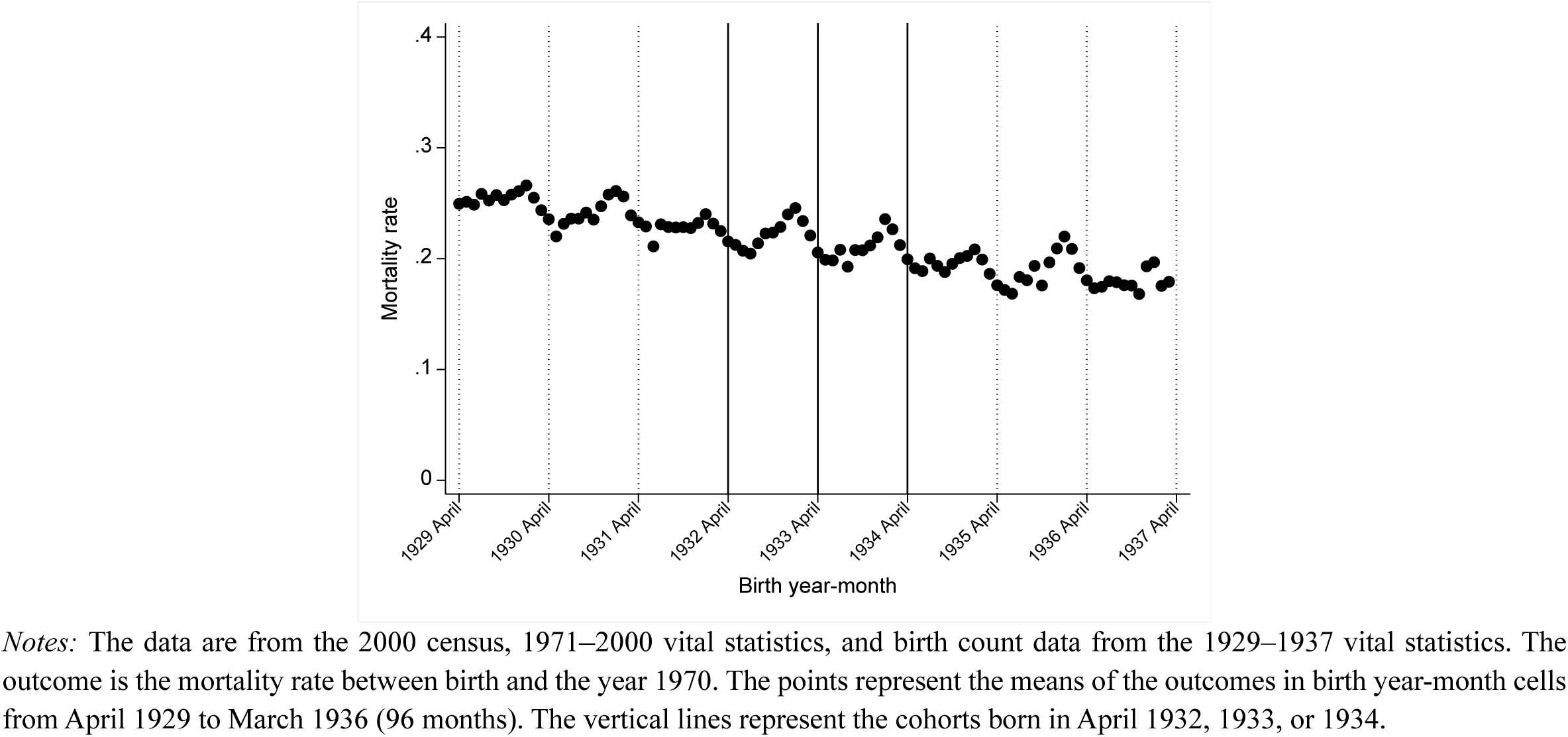
—Mortality rate between birth and 1970.

**Figure C2.**
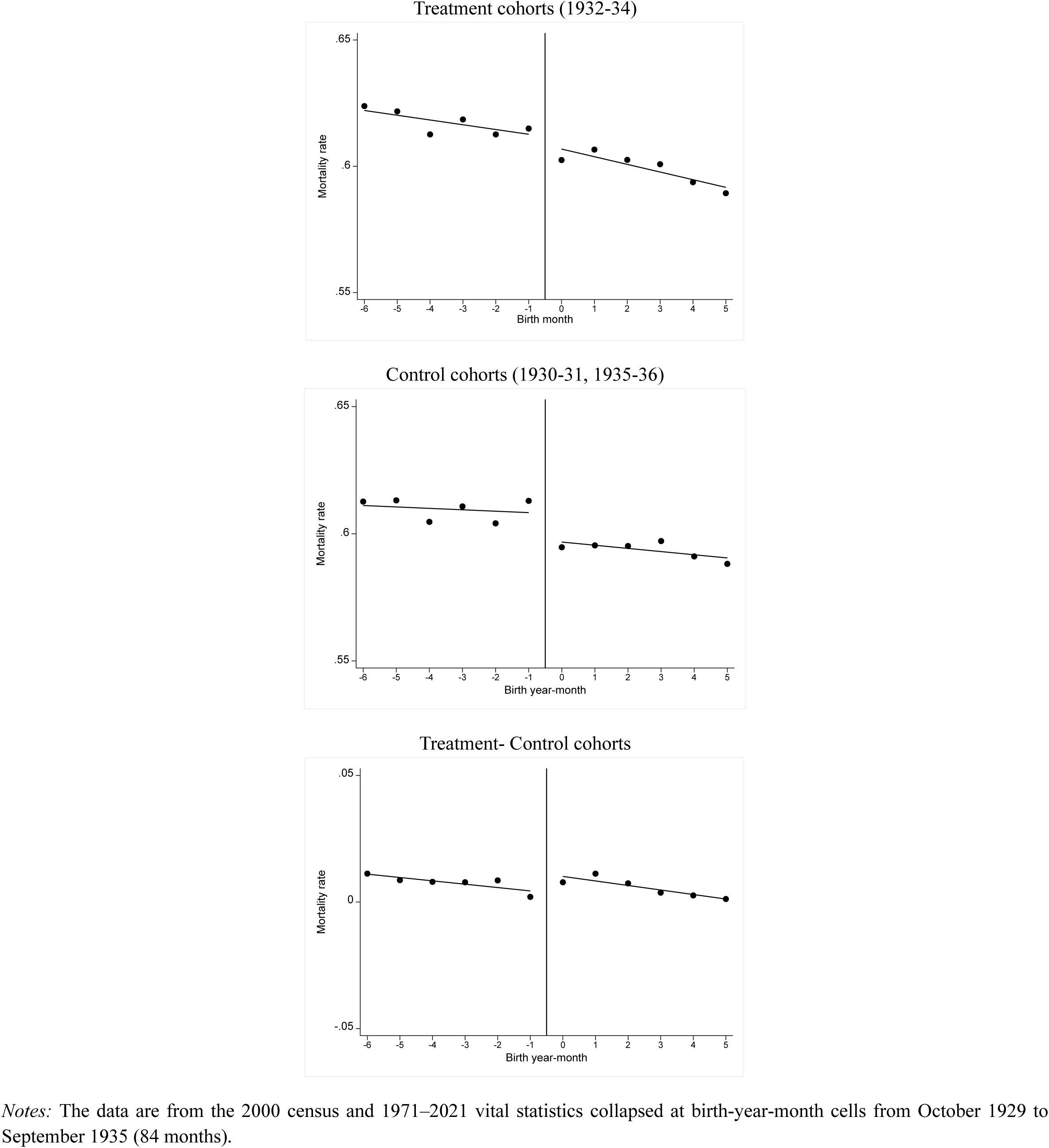
—All-cause mortality by Dif-in-RD.

**Figure C3.**
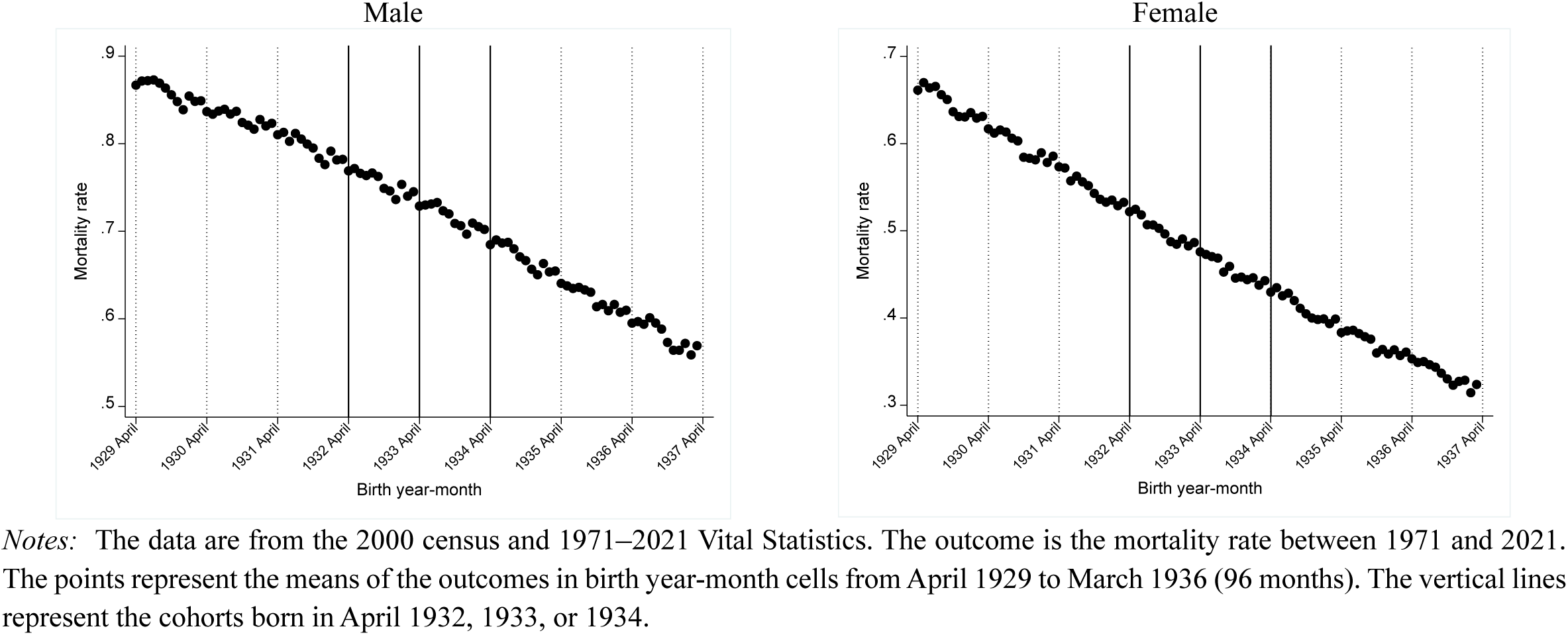
—All-cause mortality by gender.

**Table C1.** —ICD and causes of death/hospitalization.

|  | **ICD9** | **ICD10** |
| --- | --- | --- |
|  | 1971–1995 | 1996–2019 |
| **Cancer** | 140–209 | C00–C97 |
| **Circulatory diseases** | 390–459 | I00–I99 |
| **Respiratory diseases** | 460–519 | J00–J99 |
| **Treatable causes of death** |  |  |
| Tuberculosis | 010–018, 137 | A15–A19, B90 |
| Malignant neoplasm of cervix uteri | 180 | C53 |
| Chronic rheumatic heart disease | 393–398 | I05–I09 |
| All respiratory diseases | 460–519 | J00–J99 |
| Asthma | 493 | J45, J46 |
| Appendicitis | 540–543 | K35–K38 |
| Abdominal hernia | 550–553 | K40–K46 |
| Hypertensive and cerebrovascular disease | 401–405, 430–438 | I10–I15, I60–I69 |
| Cholelithiasis and cholecystitis | 574, 575.0, 575.1 | K80–K81 |
| **Preventable causes of death** |  |  |
| Lung cancer | 162 | C33–C34 |
| Cirrhosis of liver | 571.0–571.3, 571.5–571.6 | K70, K74.3–K74.6 |
| External causes of death | 800–999 | V, W, X, Y |

*Notes:* “Treatable causes of death” and “Preventable causes of death” are from Meghir et al. (2018).

**Table C2.** —All-cause mortality by gender.

|  | All causes |  |
| --- | --- | --- |
|  | Male<br>(1) | Female<br>(2) |
| **A. RD with multiple cutoffs** |  |  |
| 1932 cutoff | -0.0068***<br>(0.0024) | 0.0013<br>(0.0027) |
| 1933 cutoff | -0.0024<br>(0.0023) | 0.0004<br>(0.0029) |
| 1934 cutoff | -0.0012<br>(0.0027) | -0.0019<br>(0.0026) |
| Mean below 1932 cutoff | 0.795 | 0.546 |
| Mean below 1933 cutoff | 0.755 | 0.499 |
| Mean below 1934 cutoff | 0.715 | 0.454 |
| N of birth year-month | 96 | 96 |
| N of observations | 6,665,444 | 6,572,450 |
| **B. Dif-in-RD (pooled cutoffs)** |  |  |
| 1(After April) | -0.0119***<br>(0.0017) | -0.0096***<br>(0.0020) |
| 1(After April)×Treated | 0.0007<br>(0.0028) | 0.0045<br>(0.0030) |
| Mean | 0.745 | 0.489 |
| N of birth year-month | 84 | 84 |
| N of observations | 5,855,484 | 5,768,910 |

*Notes:* The data are from the 2000 census, and 1971–2021 vital statistics collapsed at birth year-month cells from April 1929 to March 1936 (96 months) in Panel A and October 1929 to September 1935 (84 months) in Panel B. Panel A reports the estimates from equation [1], with robust standard errors in parentheses. The 1932, 1933, and 1934 cutoffs are dummies that take 1 when the cohort is above April each year. Panel B reports the estimates from equation [2] with robust standard errors in parentheses. 1(After April) is an indicator equaling 1 for individuals born after the month of April and 0 otherwise. Treated is an indicator equaling 1 for treated cohorts born between October 1931 and September 1934 and 0 otherwise. The number of observations reflects the number of survivors up to January 1971, our population at risk. Significance levels: \*\*\* $p < 0.01$ , \*\* $p < 0.05$ , \* $p < 0.10$ .

**Table C3.** —Heterogeneity (all-cause mortality)

|  | Baseline | Period |  | Ages |  |  |
| --- | --- | --- | --- | --- | --- | --- |
|  |  | 1972–1999 | 2000–2021 | 50–59 | 60–69 | 70–79 |
|  | (1) | (2) | (3) | (4) | (5) | (6) |
| **A. RD with multiple cutoffs** |  |  |  |  |  |  |
| 1932 cutoff | -0.0025<br>(0.0025) | -0.0022**<br>(0.0010) | -0.0010<br>(0.0028) | -0.0008<br>(0.0006) | -0.0018**<br>(0.0008) | -0.0007<br>(0.0011) |
| 1933 cutoff | 0.0002<br>(0.0027) | -0.0019<br>(0.0013) | 0.0011<br>(0.0029) | -0.0001<br>(0.0007) | -0.0015<br>(0.0012) | -0.0014<br>(0.0014) |
| 1934 cutoff | 0.0005<br>(0.0038) | -0.0013<br>(0.0020) | 0.0016<br>(0.0037) | -0.0002<br>(0.0010) | -0.0013<br>(0.0012) | -0.0007<br>(0.0017) |
| Mean below 1932 cutoff | 0.672 | 0.167 | 0.606 | 0.047 | 0.093 | 0.184 |
| Mean below 1933 cutoff | 0.628 | 0.153 | 0.561 | 0.046 | 0.091 | 0.181 |
| Mean below 1934 cutoff | 0.585 | 0.142 | 0.517 | 0.045 | 0.089 | 0.178 |
| N of birth year-month | 96 | 96 | 96 | 96 | 96 | 96 |
| N of observations | 13,237,894 | 13,237,894 | 11,304,313 | 12,849,617 | 12,190,490 | 10,956,628 |
| **B. Dif RD (pooled cutoffs)** |  |  |  |  |  |  |
| 1(After April) | -0.0071***<br>(0.0016) | -0.0024***<br>(0.0008) | -0.0070***<br>(0.0016) | -0.0008**<br>(0.0003) | -0.0009*<br>(0.0005) | -0.0015<br>(0.0009) |
| 1(After April)×Treated | 0.0033<br>(0.0026) | -0.0004<br>(0.0013) | 0.0041<br>(0.0027) | 0.0001<br>(0.0006) | -0.0007<br>(0.0010) | 0.0011<br>(0.0014) |
| Mean below cutoff | 0.617 | 0.150 | 0.550 | 0.046 | 0.090 | 0.179 |
| N of birth year-month | 84 | 84 | 84 | 84 | 84 | 84 |
| N of observations | 11,624,394 | 11,624,394 | 9,917,401 | 11,281,888 | 10,700,457 | 9,612,811 |

Notes: The data are from the 2000 census, and 1971–2021 vital statistics collapsed at birth year-month cells from April 1929 to March 1936 (96 months) in Panel A and October 1929 to September 1935 (84 months) in Panel B. Panel A reports the estimates from equation [1], with robust standard errors in parentheses. The 1932, 1933, and 1934 cutoffs are dummies that take 1 when the cohort is above April each year. Panel B reports the estimates from equation [2] with robust standard errors in parentheses. 1(After April) is an indicator equaling 1 for individuals born after the month of April and 0 otherwise. Treated is an indicator equaling 1 for treated cohorts born between October 1931 and September 1934 and 0 otherwise. The number of observations reflects the number of survivors up to January 1971, our population at risk. Significance levels: \*\*\* p<0.01, \*\* p<0.05, \* p<0.10.

## Appendix D: Results on healthcare utilization

**Figure D1.**
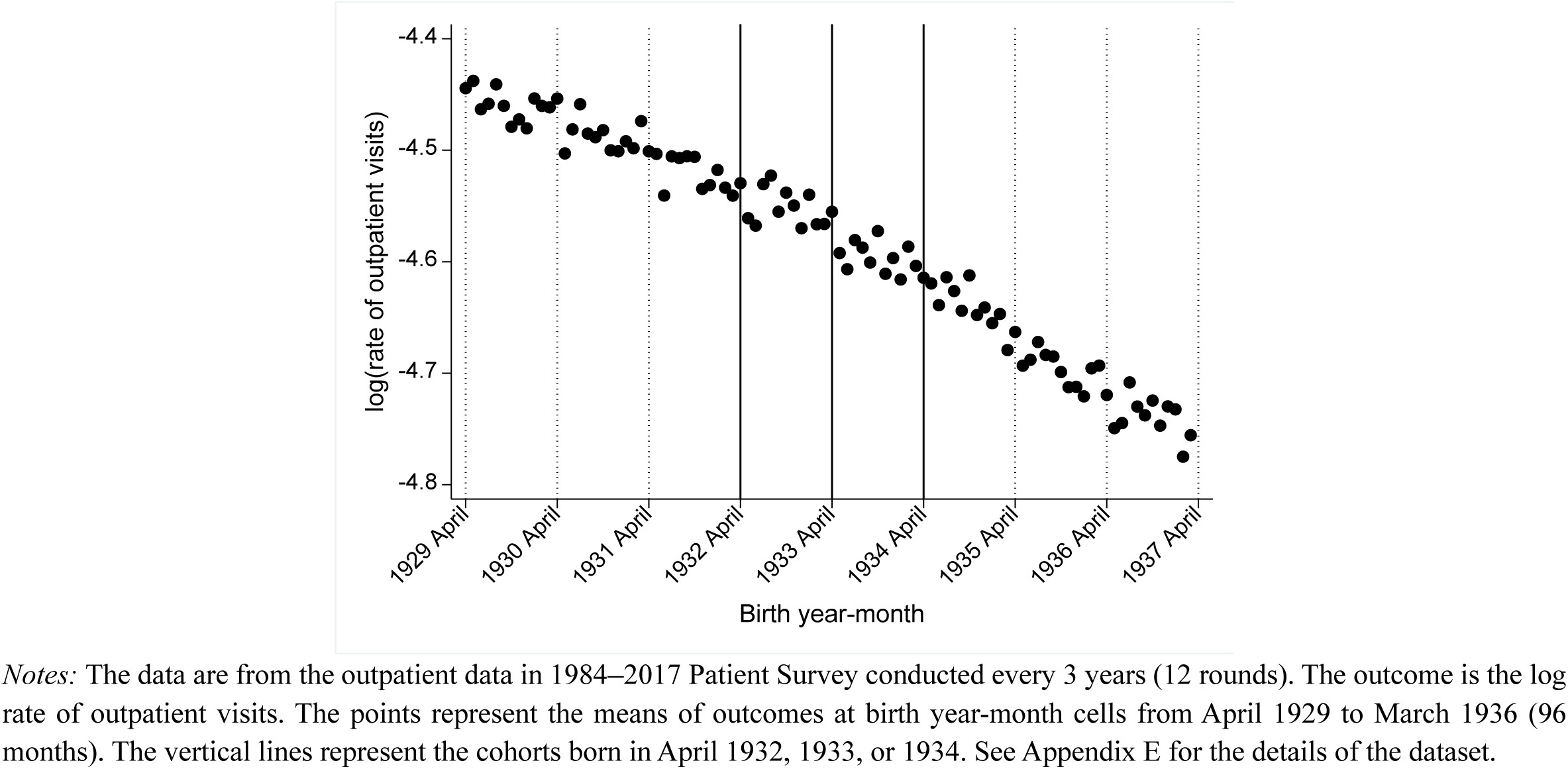
—Outpatient care.

**Figure D2.**
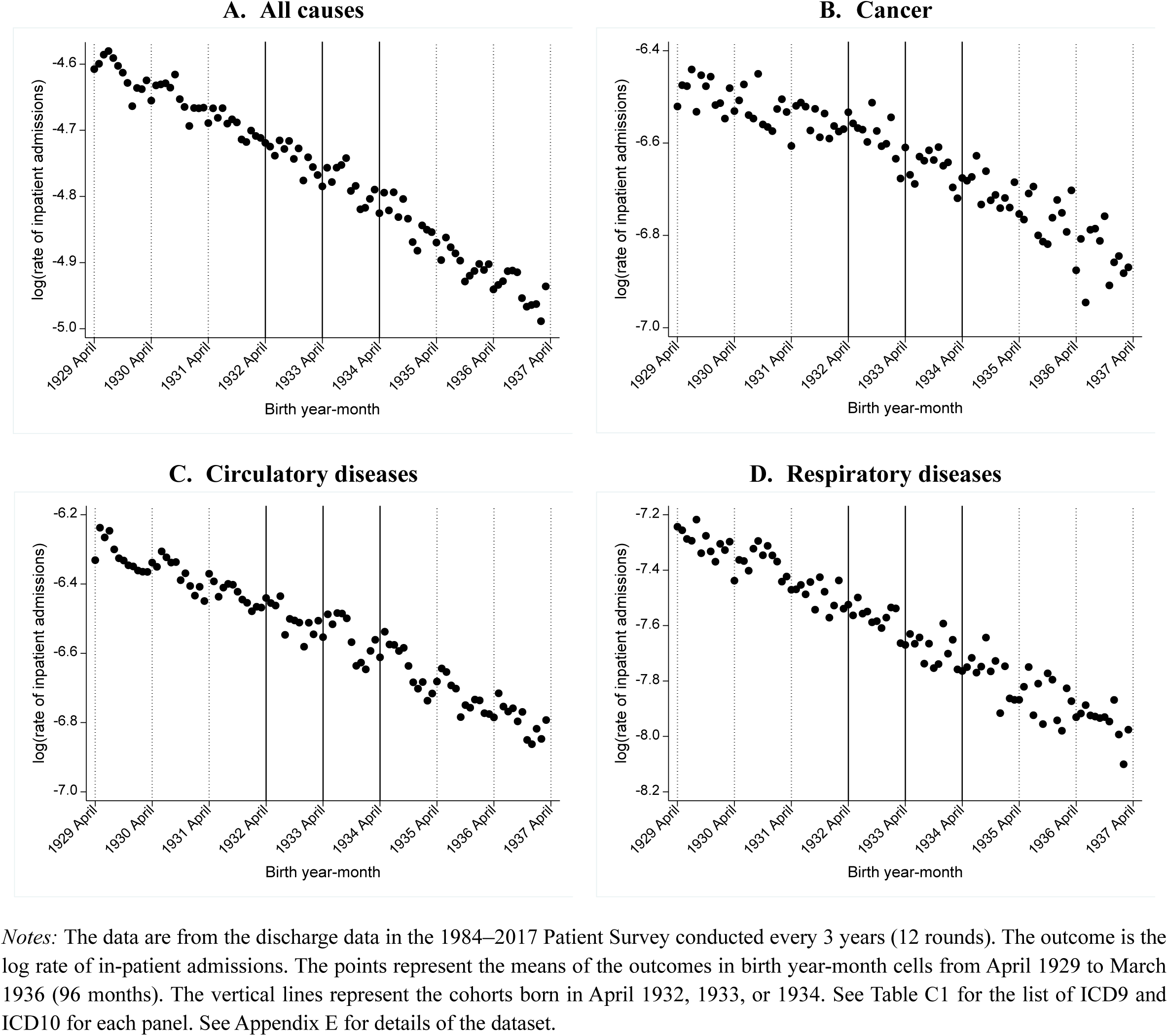
—Inpatient care (hospitalization)

**Table D1.** —Healthcare utilization.

|  | Outpatient<br>care | Inpatient care |  |  |  |
| --- | --- | --- | --- | --- | --- |
|  |  | All causes | Cancer | Circulatory<br>diseases | Respiratory<br>diseases |
|  | (1) | (2) | (3) | (4) | (5) |
| **A. RD with multiple cutoffs** |  |  |  |  |  |
| 1932 cutoff | -0.004<br>(0.012) | -0.007<br>(0.011) | 0.040<br>(0.038) | -0.023<br>(0.024) | 0.021<br>(0.039) |
| 1933 cutoff | -0.014<br>(0.015) | -0.006<br>(0.014) | 0.010<br>(0.046) | 0.001<br>(0.030) | -0.010<br>(0.052) |
| 1934 cutoff | 0.009<br>(0.018) | 0.005<br>(0.016) | 0.042<br>(0.046) | -0.005<br>(0.034) | -0.040<br>(0.063) |
| N of birth year-month | 1,152 | 1,152 | 1,152 | 1,152 | 1,152 |
| **B. Dif RD (pooled cutoffs)** |  |  |  |  |  |
| 1(After April) | -0.018<br>(0.071) | -0.018<br>(0.163) | -0.058<br>(0.225) | 0.059<br>(0.184) | -0.003<br>(0.203) |
| 1(After April)×Treated | 0.013<br>(0.108) | 0.008<br>(0.250) | 0.084<br>(0.344) | -0.035<br>(0.276) | -0.037<br>(0.313) |
| N of birth year-month | 1,008 | 1,008 | 1,008 | 1,008 | 1,008 |

*Notes:* The data are from outpatient data (column 1) and discharge data (columns 2–5) in the 1984–2017 Patient Survey conducted every 3 years (12 rounds). The outcome is the log rate of outpatient visits (column 1) and in-patient admissions (columns 2–5). The data are collapsed at birth year-month cells for each survey round from April 1929 to March 1936 (96 months) in Panel A and October 1929 to September 1935 (84 months) in Panel B. Panel A reports the estimates from an equation [1] with robust standard errors in parenthesis. The 1932, 1933, and 1934 cutoffs are dummies that take 1 when the cohort is above April each year. Panel B reports the estimates from equation [2], with standard errors clustered at birth year-month in parentheses. 1(After April) is an indicator equaling 1 for individuals born after the month of April and 0 otherwise. Treated is an indicator equaling 1 for treated cohorts born between October 1931 and September 1934 and 0 otherwise. See Table C1 for a list of ICD9 and ICD10 for each cause of death. See Appendix E for details of the dataset. Significance levels: \*\*\* p<0.01, \*\* p<0.05, \* p<0.10.

## Appendix E: Data Appendix

### 1. Social Stratification and Social Mobility Survey (SSM)

*Source:*

https://ssjda.iss.u-tokyo.ac.jp/Direct/gaiyo.php?eid=0762&lang=eng (1985)

https://ssjda.iss.u-tokyo.ac.jp/Direct/gaiyo.php?eid=0763&lang=eng (1995)

The nationwide survey on social stratification and social mobility, also known as the SSM Survey, is one of the most traditional large-scale social surveys in Japan, and has been carried out every 10 years since the first survey in 1955 by the Japan Sociological Society. It covers around 2,000 individuals residing in Japan per survey round. Individual-level data for male samples are available from 1955, and that for females first became available in 1985. Information on the individual’s highest level of education under either the old or new education system, along with information on birth year (not birth year-month), is available only for 1995 or earlier. Therefore, we use 1985 and 1995 surveys to compute the years of schooling for secondary school completion at the birth-year level and assign it to each academic cohort (allowing for measurement error), as shown in Table A1.

### 2. Patient Survey

*Source:*

http://www.mhlw.go.jp/english/database/db-hss/dl/sps_2008_06.pdf

Started in 1948, the Patient Survey is a national sample survey of hospitals and clinics that gathers information on the utilization of medical institutions in Japan. A comprehensive version of the Patient Survey has been conducted by the Ministry of Health, Labour and Welfare every 3 years since 1984. It covers roughly 2,000–7,000 hospitals and 3,000–6,000 clinics per survey year. The sample size has become larger in recent years. The survey collects information on the International Classification of Diseases (ICD) codes; patients’ principal sources of payment; and limited sociodemographic characteristics, such as gender and patients’ place of residence. Individual-level patient microdata files are available from 1984. There are two datasets in the Patient Survey, namely, 1) outpatient data and 2) discharge data, which we use to examine outpatient visits and inpatient admissions, respectively. The outpatient data in the Patient Survey are collected on a day in mid-October (normally a weekday in the second week) and include information on all patients who visit hospitals or clinics as outpatients (i.e., visits to hospitals or clinics not culminating in hospitalization). The datasets contain 364,000–1,020,000 outpatient visitors. The discharge data in the Patient Survey report all the inpatients discharged from the surveyed hospitals and clinics in the month of September of the survey year. The datasets contain about 180,000–1,140,000 inpatient records per survey year.

## Footnotes

* The authors thank Yutaka Arimoto, Anna Nicińska, and participants at the Essen Health Conference 2023 for suggestions, and Ryuich Tanaka for comments at the early stage of the project. Masuda acknowledges financial support from JSPS KAKENHI (22H00847, 19K13677). Shigeoka acknowledges financial support from JSPS KAKENHI (23H00828, 22H00057, 22H00847, 22H05009).

† Department of Economics, Gakushuin University, Toshima, Tokyo, Japan, and Hitotsubashi University.

‡ Department of Economics, Simon Fraser University, Burnaby, BC, Canada, The University of Tokyo, IZA, and NBER.

1 See studies for the US (Lleras-Muney 2005; Mazumder 2008), the UK (Clark and Royer 2013; Davies et al. 2018), France (Albouy and Lequien 2009), the Netherlands (Van Kippersluis et al. 2011), Sweden (Meghir et al. 2018; Lager and Torssander 2012; Fischer et al. 2021), Romania (Malamud et al. 2023), and Taiwan (Kan 2016). Grossman (2006), Mazumder (2012), and Galama et al. (2018) provide reviews of the literature.

2 For example, Barcellos et al. (2021) find that while the effect of a 1972 UK compulsory schooling reform on body mass index (BMI) is close to zero on average, the effect is larger at the bottom of the BMI distribution.

3 Van Kippersluis et al. (2011) examine the effect of a 1928 Dutch reform that raised the compulsory years of schooling from 6 to 7 on mortality between the ages of 81 and 87 years, but a limitation of their study is the selective mortality before age 81 years. By contrast, we observe mortality of affected cohorts from the ages of 37 to 87 years.

4 Households that do not send their children for compulsory education first get a warning from the government, and then are charged a fine of 1000 Yen as a penalty for non-compliance by law (Cabinet Office 1947).

5 The men affected by the reform were too young to fight in the war.

6 Up to the 2010 census, the lowest category was “primary/secondary school,” which combined primary and secondary school.

7 We are aware of two surveys (the National Survey for Mobility and Time Use Survey of Japan, for 2016 and 2021) which also distinguish between primary and secondary schools. However, the sample sizes of these surveys are too small to detect any discernable impact of the 1947 school reform on educational attainment.

8 See Appendix E for details of the dataset. For those who have completed high school or above, we do not know whether they went to either the old 2-year or new 3-year secondary schools before advancing to high school. To be conservative, we uniformly assign 12, 14, 16, and 18 years for those who completed high school, junior college, college, and graduate education, respectively. To the extent that recent cohorts who have completed high school or above attend a new secondary school, our estimate of the years of schooling provides the lower bound.

9 A similar approach to computing mortality is taken by Clark and Royer (2013) for the UK and Malamud et al. (2023) for Romania.

10 Following Lleras-Muney (2005), we also calculate the cohort mortality rate by the change in the cohort size between the 2020 census and the 2000 census, the first census with birth month information available, and similar results are obtained (not shown).

11 Our results are quantitatively similar when we include two or four cohorts (instead of three) before and after the school reform (not shown).

12 Specifically, it takes 1 for three treated “synthetic” cohorts, namely, those born between October 1932 and September 1933, October 1933 and September 1934, and October 1934 and September 1935, and 0 otherwise. In practice, *treat_*b*_* dummy is replaced by each “synthetic” cohort dummy to be more flexible.

13 Our results are robust to using the data only *after* the school reform as the control cohorts (not shown). We choose the current approach to keep the samples for estimating equations [1] and [2] as close as possible.

14 Panel A of Table B1 shows that the fraction of those who completed secondary school or higher increases by 2.5–3.6 and 2.3–3.9 percentage points for men and women, respectively, which translates into 0.16–0.22 and 0.09–0.19 additional years of schooling for men and women, respectively.

15 The high-skill dummy is defined as those who work as professionals or managers, following International Labour Organization standards.

